# Developmental pathways from childhood neurodevelopmental traits to early adolescent psychiatric dimensions: the role of environmental and lifestyle factors

**DOI:** 10.1101/2025.04.29.25326578

**Authors:** Chiara Caserini, Divyangana Rakesh, Shiqi Lu, Rachael Bedford, Cathryn M. Lewis, Margherita Malanchini, Giorgia Michelini

## Abstract

**Objectives:** Transdiagnostic dimensional approaches have advanced our understanding of psychiatric comorbidity and developmental continuity, but have rarely been applied to investigate links between neurodevelopmental (e.g., attention-deficit/hyperactivity disorder, autism) and other psychiatric conditions. Building on recent research delineating a transdiagnostic “neurodevelopmental spectrum”, we examined longitudinal associations between this spectrum in late childhood and other psychiatric dimensions in early adolescence, and the role of environmental/lifestyle factors as potential mediators of these associations.

**Method:** In 11,875 children from the Adolescent Brain Cognitive Development (ABCD) study, we extracted neurodevelopmental, externalizing, internalizing, detachment, and somatoform dimensions at ages 10, 11, and 12 years through confirmatory factor analysis of Child Behavior Checklist items. Using linear mixed models, we tested prospective associations between the neurodevelopmental spectrum at age 10 and other psychiatric dimensions at ages 11 and 12. Mediation models examined whether environmental (e.g., family conflict, school involvement) and lifestyle (e.g., sleep problems, screen use) factors at age 11 mediated these associations.

**Results:** Strong longitudinal associations emerged between the neurodevelopmental spectrum and all other psychiatric dimensions (β=0.417-0.641, p<0.001). Sleep problems were the most consistent mediator, accounting for 12-29% of the associations with all outcomes. Most results remained significant after multiple testing corrections and adjusting for covariates, mediators, and outcomes at baseline.

**Conclusion:** Our transdiagnostic dimensional approach revealed a wide range of poor dimensional psychiatric outcomes in children with high neurodevelopmental traits. Sleep problems emerged as a key lifestyle factor underlying these associations and promising target for transdiagnostic prevention programs to improve mental health outcomes in this population.

## INTRODUCTION

Neurodevelopmental conditions (NDCs), including attention-deficit/hyperactivity disorder (ADHD), autism, and motor, language, and learning disorders, jointly affect around 15% of children and young people worldwide^1,2^, with many more showing subthreshold traits. NDCs are classified as separate “neurodevelopmental disorder” categories in current diagnostic systems^3,4^, and are often assessed and supported by different clinical services and professionals. However, this classification does not align with evidence showing their frequent co-occurrence^5–7^ and substantial overlap in clinical features and underpinnings^8^. Moreover, children and young people with NDCs are at substantially elevated risk for other mental health conditions relative to their neurotypical peers^9,10^. Externalizing and internalizing problems are especially common^9,10^, with emerging evidence further suggesting increased risk for psychosis^11^. These mental health comorbidities and their early manifestations (e.g., psychotic traits) often emerge in early adolescence, with widespread negative impacts on education, social relationships, and quality of life^12^. However, children and young people with NDCs show limited benefit from pharmacological and psychological mental health treatments developed in neurotypical samples^12^, potentially suggesting different underlying mechanisms. As mental health is a top priority of individuals with NDCs and their families globally^13,14^, new approaches are needed to understand the processes underlying mental health outcomes in this population and design targeted interventions.

Most studies of mental health outcomes in NDCs have focused on individual diagnoses, mainly autism or ADHD^15,16^. This approach does not account for the high rates of NDC co-occurence, the many overlapping characteristics across diagnoses, and the continuous distribution of NDC traits in the population^10^. Moreover, different NDCs are associated with a similar increase in the risk for mental health conditions, both as categorical diagnoses and dimensional traits^9,17,18^. These findings have recently motivated researchers to apply transdiagnostic and dimensional approaches^19,20^ spanning traits of all NDC diagnoses included in categorical diagnostic manuals^3,4^ and conceptualize them as part of a broad “neurodevelopmental spectrum”^10^. This approach extends current transdiagnostic and hierarchical dimensional frameworks, such as the Hierarchical Taxonomy of Psychopathology (HiTOP)^21^. While these frameworks organize many psychiatric symptoms in well-established dimensions (e.g., externalizing, internalizing, detachment), they do not currently include traits of most NDCs, except for ADHD^10^. Focusing on a transdiagnostic neurodevelopmental spectrum alongside other psychiatric dimensions offers a useful approach for understanding the developmental overlap between NDCs and mental health conditions, particularly in large epidemiological samples including young people with traits of one or more NDCs. However, no study to date has examined the longitudinal association between the transdiagnostic neurodevelopmental spectrum and other psychiatric dimensions.

Besides charting associations over development, pinpointing the processes underlying the emergence of mental health conditions in children with NDCs is crucial to guide the development of novel mental health interventions tailored to their needs. Modifiable aspects of the environment and lifestyle that mediate links between early neurodevelopmental traits and later mental health outcomes may be particularly promising intervention targets. Initial longitudinal research in this area, focused on ADHD^15,22–24^ and autism^25–27^, suggests that lower quality relationships with family and peers, including associated negative life events such as bullying, and children’s involvement in their school environment could mediate the link between neurodevelopmental traits and poor psychiatric outcomes, such as externalizing and internalizing problems. Neighborhood characterisitcs, including safety, have further been associated with NDCs^28,29^ and shown to prospectively predict mental health in young people from the general population^30^. With regard to lifestyle factors, initial longitudinal findings suggest that sleep problems are associated with mental health outcomes in ADHD^31^ and autistic^32^ samples. Poor mental health outcomes in longitudinal population-based youth samples have also been linked with lower physical activity^33^, unhealthy dietary patterns^34,35^, and screen use (albeit with small effects)^36,37^.

However, findings on environmental and lifestyle factors have been somewhat mixed, with studies reporting both significant and null effects, and many potential mediating effects have not been tested formally in children with NDC diagnoses or traits. Many studies have also examined these factors in isolation, without providing a comprehensive examination or accounting for broader challenges (e.g., low socioeconomic status). Moreover, few studies have employed rigorous longitudinal mediation designs, which are essential for inferring directionality and identifying intervention targets^38^. Finally, no study has examined environmental/lifestyle factors underlying risk for a range of psychiatric outcomes in children with transdiagnostic neurodevelopmental traits, which can inform future intervention strategies with far-reaching effects.

The current study sought to address these research gaps using three waves of longitudinal data from almost 12,000 children in the Adolescent Brain Cognitive Development (ABCD) study. First, we comprehensively investigated to what extent the neurodevelopmental spectrum in childhood prospectively predicts major dimensions of psychiatric symptoms previously identified in this cohort^20^ across two time points spanning early adolescence. Specifically, we considered externalizing and internalizing dimensions, extending previous literature on individual NDCs, but also detachment (early manifestations of negative psychotic symptoms, e.g., social withdrawal, low energy) and somatoform (somatic complaints), which have been scarcely investigated in association with NDCs. Second, we tested the role of putative environmental and lifestyle factors (parental warmth, parental monitoring, family conflict, school involvement, negative life events, neighborhood safety, sleep problems, physical activity, diet, screen use) as mediators of these longitudinal associations, selected based on the extant literature. We hypothesized that children with higher scores on the neurodevelopmental spectrum would display higher levels of psychopathology across all investigated dimensions over time, and that environmental and lifestyle factors would partly mediate these associations, pointing to promising intervention targets.

## METHODS

### Sample

Our study included 11,875 participants from the Adolescent Brain Cognitive Development (ABCD) study, one of the largest longitudinal epidemiological studies investigating brain and behavioral development across adolescence^39^. The ABCD sample recruited children aged 9-10 years from schools across 21 sites in the US with the aim to collect a diverse sample in terms of gender, race, ethnicity, socioeconomic status, and urbanicity (for complete recruitment details see^39^). As a result, the sample approaches the diversity of the US population on several socio-demographic characteristics. For example, 51% of participants are White, 21% are Hispanic, 15% are African American, 2% are Asian, and 10% are multiracial or from other ethnical backgrounds. Household income at baseline was <$50,000 for 31% of families, between $50,000 and <$100,000 for 28% of families, and at least $100,000 for 41% of families; 59% of children had at least one parent with a bachelor’s or postgraduate degree. Institutional Review Board approval was obtained at the University of California, San Diego, and in some cases at individual sites. All parents/guardians provided written informed consent and children provided assent before participation in the study.

The current longitudinal analyses used data from the baseline (N=11,875, % males=52, mean age=9.914, SD=0.005; “age 10” henceforth), 1-year follow up (N=11,219, % males= 52, mean age=10.923, SD=0.006; “age 11”), and 2-year follow up (N=10,972, % males=52, mean age=12.026, SD=0.006; “age 12”) from the ABCD 5.1 data release (DOI: 10.15154/z563-zd24).

### Measures

We measured dimensions of neurodevelopmental traits and mental health symptoms at the three time points (ages 10 to 12) and environmental and lifestyle factors at ages 10 and 11.

#### Neurodevelopmental traits and mental health symptoms

Parents completed the Child Behavior Checklist 6-18 (CBCL/6-18)^40^, which was used to extract the neurodevelopmental, internalizing, externalizing, somatoform, and detachment factors at each wave (see “Statistical analyses”), based on the factor structure previously identified in this sample at age 10^20^.

#### Environmental and lifestyle factors

We considered a broad range of environmental and lifestyle factors as candidate mediators of the relationship between neurodevelopmental traits and later mental health, guided by previous literature^15,22–37^. Full details on measures are presented in Supplement 1. Briefly, environmental factors included measures of child-reported parental warmth, parental monitoring, family conflict, school involvement and feelings about school, number of negative life events, and neighborhood safety. Lifestyle factors included measures of child-reported sleep problems and parent-reported duration of screen use (distinguishing between social media, social engagement, passive watching, and active gaming), involvement in physical activities and sports, and healthy dietary patterns during a typical week. All the environment and lifestyle measures were available at ages 10 and 11, except for dietary patterns and negative life events, which were collected only at age 11.

### Statistical analyses

All analysis code for MPlus and R is available at: https://osf.io/yhkvm/.

Before testing our study aims, we used Confirmatory Factor Analyses (CFA) in Mplus 8^41^ to extract five transdiagnostic factors of neurodevelopmental traits and mental health problems previously identified by Michelini et al.^20^ in the ABCD study at baseline (age 10) from the CBCL items. Consistent with Michelini et al.^20^, we removed items with very low endorsement (>99.5% rated 0) and aggregated items that were highly correlated (polychoric r>0.75) into composites before CFA. Analyses included all available participants, including twins and siblings (excluded in Michelini et al.^20^), by clustering within family using “CLUSTER” and “TYPE = COMPLEX” commands. Items without a clear primary loading or cross-loading items in Michelini et al.^20^ were not considered, leaving 69 items. Based on the primary loadings on the specific factors^20^, we extracted neurodevelopmental, externalizing, internalizing, detachment, and somatoform factors (Figure 1). The neurodevelopmental factor primarily included inattention and hyperactivity, but also cognitive disengagement, motor difficulties, autistic-like traits, atypical ideation, and poor school performance. The externalizing factor captured aggressive, oppositional, and rule-breaking behaviors. The internalizing factor included symptoms of low mood, anxiety, and phobias. The detachment factor captured social withdrawal, shyness, and lack of energy. Finally, the somatoform factor included somatic complaints and pain.

**Figure 1.**
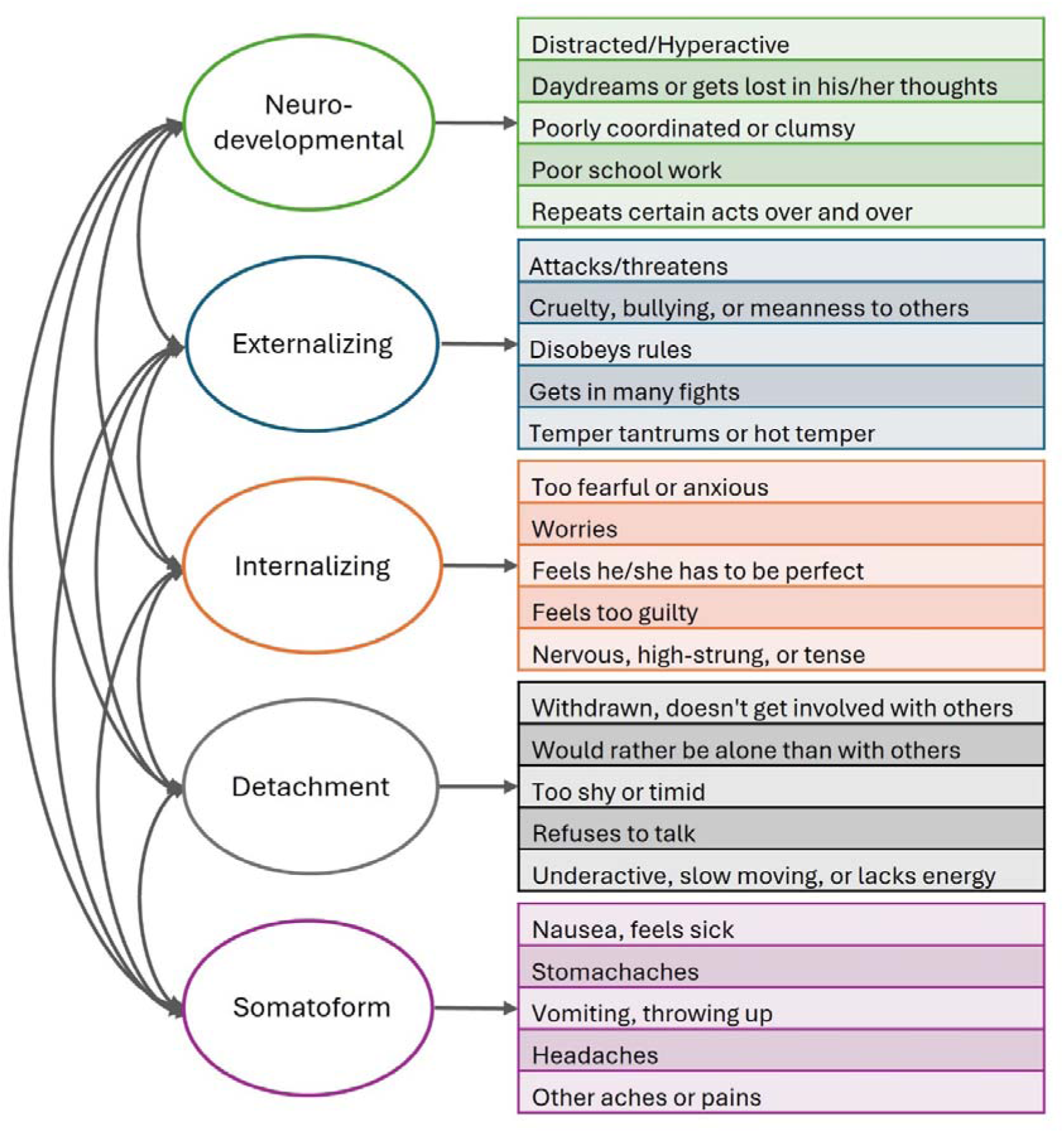
Graphical representation of the correlated factors model of Child Behavior Checklist (CBCL) items based on Michelini et al. (2019)^20^, including examples of items with significant primary loadings.

To investigate the longitudinal link between the neurodevelopmental factor and mental health outcomes (first aim), linear mixed models tested the association between the age-10 neurodevelopmental factor and each of the mental health factors (externalizing, internalizing, detachment, somatoform) at ages 11 and 12, accounting for family relatedness^42^. Unadjusted analyses were run on standardized residuals of sex and age to control for these covariates. Adjusted analyses further controlled for the respective mental health factors at age 10 to examine associations beyond baseline mental health problems (e.g., we accounted for age-10 internalizing factor when testing the link between age-10 neurodevelopmental factor and age-11 internalizing factor).

To identify mediators of the link between early neurodevelopmental traits and later mental health outcomes (second aim), we ran single-mediation analyses, using the “*lavaan*” package in R and bootstrap estimation to obtain robust standard errors for the estimated parameters. We estimated the direct effect of the neurodevelopmental factor on mental health outcomes without considering the mediator (path c), the effect of the neurodevelopmental factor on each environmental/lifestyle factor (path a), the effect of the environmental/lifestyle factor on mental health factors (path b), the indirect effect of each environmental/lifestyle factor in the link between the ND factor and mental health outcomes (path c’), and the proportion of total effect (c+c’) explained by the mediator (prop. mediated). These analyses used only one randomly selected child per family when more than one participated (N=9,851), as mediation models in the full sample clustering within family did not converge, likely because most participants were unrelated. In unadjusted analyses, we tested the mediation effect of each environmental/lifestyle factor at age 11 in the association between the neurodevelopmental factor at age 10 and mental health outcomes at age 11 and 12 separately, using standardized residuals of sex and age to control for these covariates. Adjusted analyses further controlled for age-10 measures of each mental health outcome and environmental/lifestyle mediator (except diet and negative life events, which were only available at age 11), to examine effects beyond baseline measures. The latter analysis using age 12 mental health outcomes provides a stronger test of potentially causal effects^38^. Multiple testing corrections were applied separately for each outcome using the Benjamini-Hochberg false discovery rate (FDR) procedure^43^ (q=0.050).

To rule out the potential effects of socioeconomic status and general cognitive ability on the identified mediation effects, sensitivity analyses further controlled for age-10 total combined family income and IQ (measured with Matrix Reasoning Total Scaled Score from the Wechsler Intelligence Test for Children-V)^44^ in adjusted models of age-12 outcomes that survived multiple testing corrections. Further, while the focus of our study was on specific factors (neurodevelopmental, externalizing, internalizing, detachment, and somatoform) from Michelini et al.^20^, for completeness we run additional analyses on the general psychopathology (‘p’) factor from this previous study.

## RESULTS

### Longitudinal associations between neurodevelopmental factor and mental health outcomes

Descriptive statistics for the neurodevelopmental and mental health factors at each time point are in Table S1. The neurodevelopmental factor at age 10 showed positive significant associations with all mental health outcomes at age 11 and 12 in unadjusted analyses (Table 1). The strongest effect was observed for the externalizing factor at age 11, where the neurodevelopmental factor explained 41% of the variance. Effects remained significant in adjusted models controlling for each of the mental health factors at age 10, showing that the neurodevelopmental factor uniquely explained approximately 10-16% of variance in outcomes after accounting for age-10 factors (Table 1).

**Table 1.** Longitudinal associations between the neurodevelopmental factor at baseline (age 10) and mental health outcomes at follow-up 1 (age 11) and follow-up 2 (age 12), in unadjusted models (controlling only for sex and age) and adjusted models (also controlling for each outcome at baseline).

|  | UNADJUSTED |  |  |  | ADJUSTED |  |  |  |  |
| --- | --- | --- | --- | --- | --- | --- | --- | --- | --- |
| Outcome | $\beta$ | SE | p | R<sup>2</sup> | $\beta$ | SE | p | R<sup>2</sup> | R<sup>2</sup> diff |
| Externalizing age 11 | 0.641 | 0.007 | <0.001 | 0.410 | 0.058 | 0.011 | <0.001 | 0.555 | 0.145 |
| Externalizing age 12 | 0.589 | 0.007 | <0.001 | 0.340 | 0.050 | 0.012 | <0.001 | 0.471 | 0.131 |
| Internalizing age 11 | 0.576 | 0.007 | <0.001 | 0.333 | 0.069 | 0.011 | <0.001 | 0.488 | 0.155 |
| Internalizing age 12 | 0.527 | 0.008 | <0.001 | 0.277 | 0.065 | 0.012 | <0.001 | 0.411 | 0.134 |
| Detachment age 11 | 0.593 | 0.007 | <0.001 | 0.352 | 0.123 | 0.011 | <0.001 | 0.469 | 0.117 |
| Detachment age 12 | 0.539 | 0.008 | <0.001 | 0.289 | 0.118 | 0.012 | <0.001 | 0.385 | 0.096 |
| Somatoform age 11 | 0.428 | 0.008 | <0.001 | 0.183 | 0.133 | 0.009 | <0.001 | 0.344 | 0.161 |
| Somatoform age 12 | 0.417 | 0.008 | <0.001 | 0.173 | 0.158 | 0.010 | <0.001 | 0.301 | 0.128 |
| Neurodevelopmental age 11 | 0.744 | 0.006 | <0.001 | 0.553 | - | - | - | - | - |
| Neurodevelopmental age 12 | 0.688 | 0.007 | <0.001 | 0.471 | - | - | - | - | - |

Note: The association between the neurodevelopmental factor at baseline (age 10) and the follow-up assessments (ages 11 and 12) is also reported at the bottom of the table for completeness.
Abbreviations: $\beta$ , beta regression coefficients; R<sup>2</sup>, variance explained; R<sup>2</sup> diff, difference in variance explained between unadjusted and adjusted models; SE, standard error.

### Mediation effects

#### Overview

In both unadjusted and adjusted analyses, sleep problems emerged as the mediator explaining the largest amount of variance (12-29%) in the association between the neurodevelopmental factor and externalizing, internalizing, detachment, and somatoform outcomes at age 11 and 12 (Figures 2 and 3, Tables S2 and S3).

**Figure 2.**
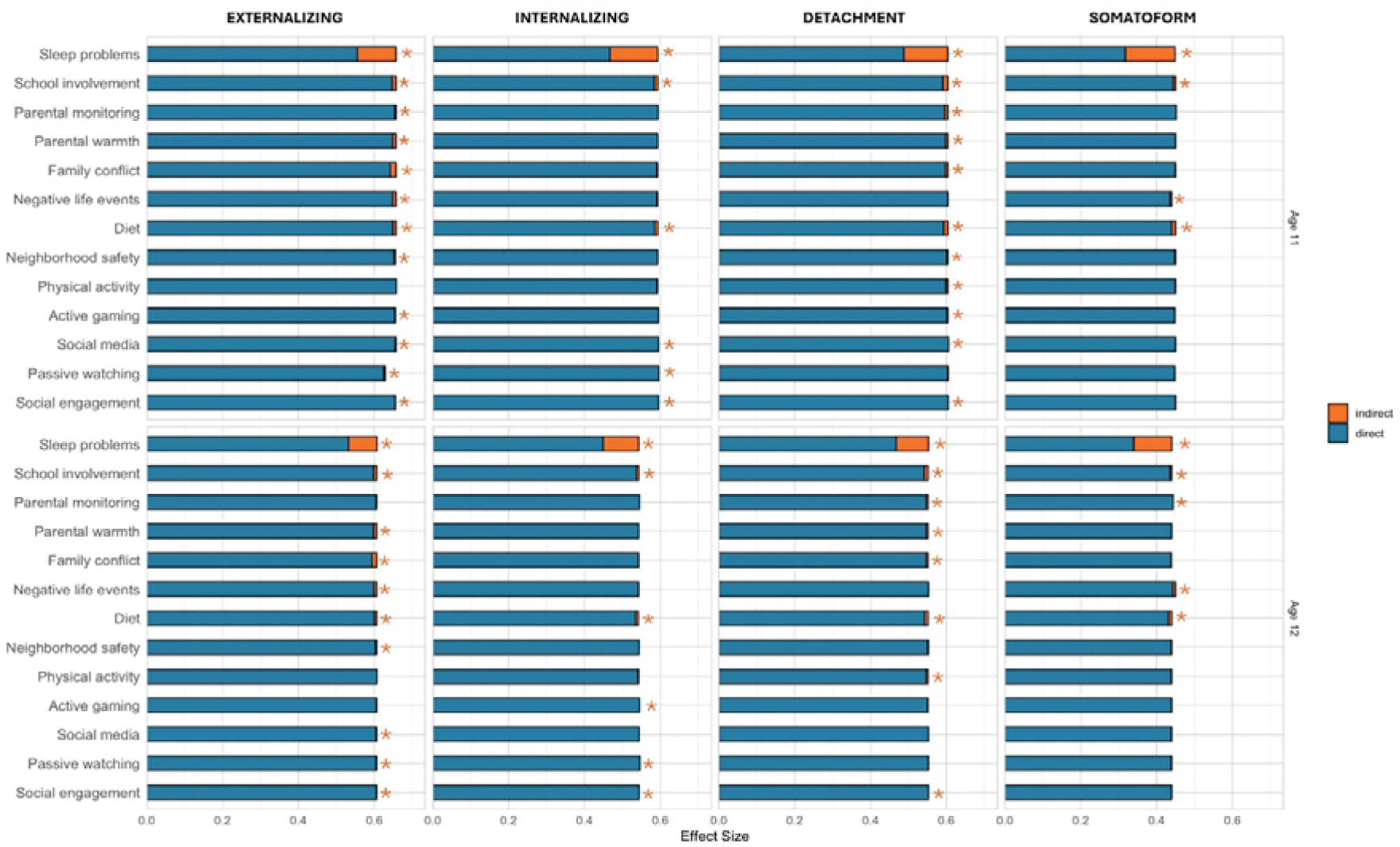
Effects of candidate mediators in the longitudinal association between the childhood neurodevelopmental spectrum and the early adolescent mental health outcomes in unadjusted models (only controlling for age and sex). Notes: The total length of each bar represents the prediction (standardised beta coefficient) from the neurodevelopmental spectrum at age 10 to each mental health outcome at ages 11 and 12. The blue portion of each bar shows the direct effect, while the orange portion shows the indirect (i.e., mediated) effect. *indicates statistically significant (p<0.05) indirect effects after applying FDR multiple testing correction.

**Figure 3.**
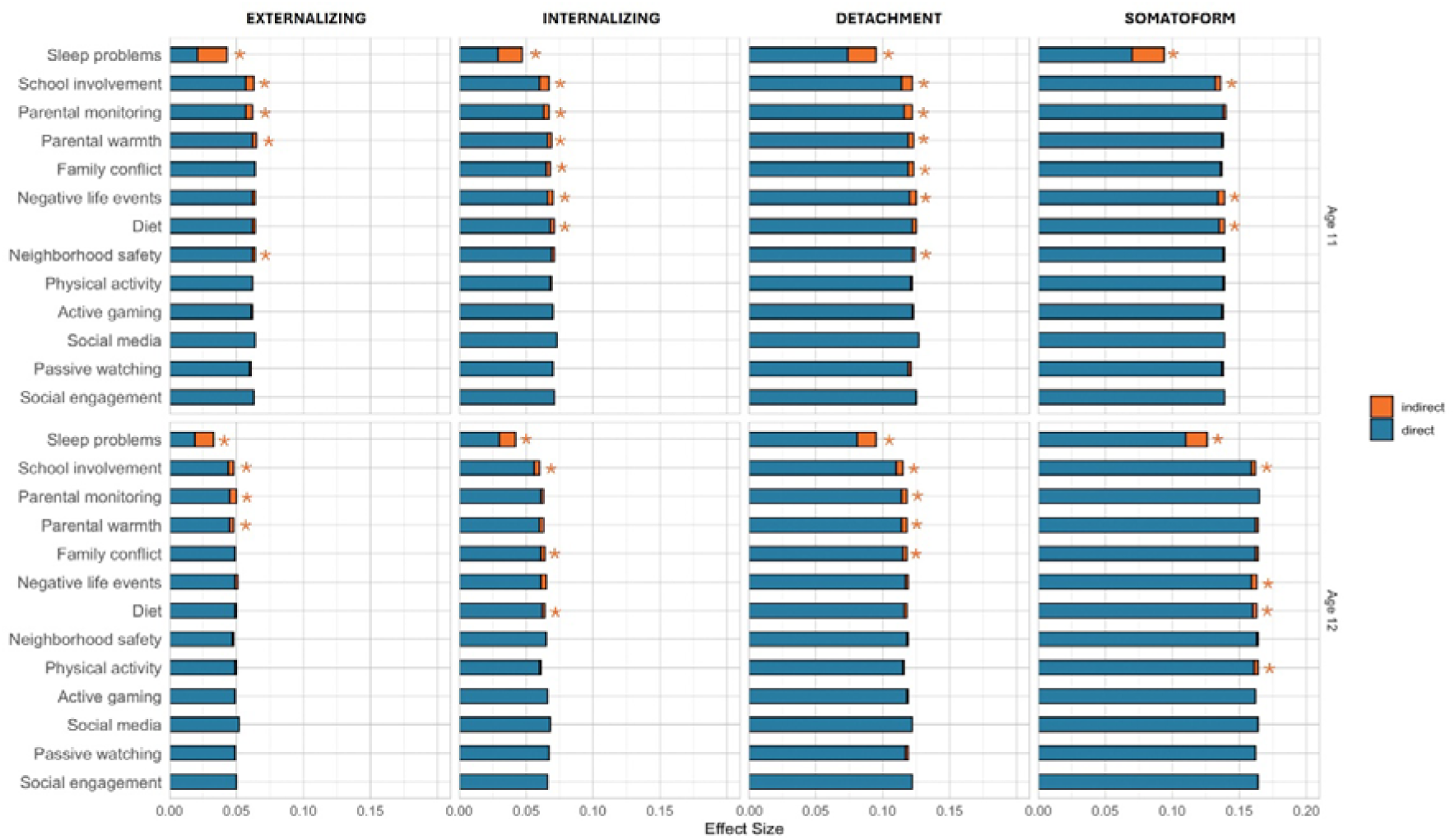
Effects of candidate mediators in the longitudinal association between the childhood neurodevelopmental spectrum and the early adolescent mental health outcomes in adjusted models (controlling for age, sex, and baseline measures of each mediator and outcome). Notes: The total length of each bar represents the prediction (standardised beta coefficient) from the neurodevelopmental spectrum at age 10 to each mental health outcome at ages 11 and 12. The blue portion of each bar shows the direct effect, while the orange portion shows the indirect (i.e., mediated) effect. *indicates statistically significant (p<0.05) indirect effects after applying FDR multiple testing correction.

#### Externalizing factor

In unadjusted mediation models (Table S2, Figure 2), each of the environmental and lifestyle factors (lower parental monitoring, parental warmth, school involvement, neighborhood safety; higher family conflict, negative life events, sleep problems, and screen use; and less healthy diet) with the exception of physical activity, significantly mediated the association between the neurodevelopmental factor at age 10 and mental health outcomes at age 11 (indirect effects: β between 0.001 and 0.102, FDR-p<0.048). All environmental and lifestyle factors, except physical activity, active gaming, and parental monitoring, were also significant mediators of outcomes at age 12 in unadjusted analyses (β between 0.001 and 0.075, FDR-p<0.040). The proportion of total effects explained by these significant mediators was between 0.2-15% at age 11 and between 0.2-12% at age 12.

When we adjusted for the respective mediator and externalizing problems at age 10 (Table S3, Figure 3), indirect effects were significant for higher sleep problems and lower parental monitoring, parental warmth, school involvement, and neighborhood safety at age 11 (β between 0.002 and 0.022, FDR-p<.007) and for higher sleep problems and lower parental monitoring, parental warmth, and school involvement at age 12 (β between 0.003 and 0.014, FDR-p<0.016). The proportion of total effects in externalizing problems explained by these significant mediators was between 3-51% at age 11 and between 6-43% at age 12.

#### Internalizing factor

In unadjusted mediation models of internalizing problems, lower school involvement, higher sleep problems, passive watching, social media, and social engagement, and less healthy diet were significant mediators of mental health outcomes at age 11 (β between -.002 and .126, FDR-p<.019). Lower school involvement, higher sleep problems, passive watching, active gaming, and social engagement, and less healthy diet were significant mediators also of outcomes at age 12 (β between -0.002 and 0.094, FDR-p<0.046). The proportion of total effects explained by these significant mediators was between -0.3 and 21% at age 11 and between -0.3 and 17% at age 12.

In adjusted models controlling for each mediator and internalizing problems at age 10, indirect effects were significant for higher family conflict, negative life events, and sleep problems, less healthy diet, and lower parental monitoring, parental warmth, and school involvement at age 11 (β between 0.002 and 0.018, FDR-p<0.034) and for higher family conflict and sleep problems, less healthy diet, and lower school involvement at age 12 (β between 0.002 and 0.012, FDR-p<0.049). The proportion of total effects explained by these significant mediators was between 2.4-38% at age 11 and between 3.8-28% at age 12.

#### Detachment factor

In unadjusted mediation models of detachment problems, all environmental and lifestyle factors were significant mediators of mental health outcomes at age 11 in expected directions (β between -0.001 and 0.116, FDR-p<0.028), except for negative life events and passive watching. All mediators except negative life events, passive watching, active gaming, and social media were also significant at age 12 (β between -0.001 and 0.085, FDR-p<0. 045). The proportion of total effects explained by these significant mediators was between 1.3-19% at age 11, and between -0.2 and 15% at age 12.

In adjusted models controlling for each mediator and detachment problems at age 10, indirect effects were significant for higher family conflict, negative life events, and sleep problems, and lower parental monitoring, parental warmth, neighborhood safety, and school involvement at age 11 (β between 0.002 and 0.021, FDR-p<.004). The same mediators, except negative life events and neighborhood safety, also showed significant indirect effects on outcomes at age 12 (β between 0.003 and 0.014, FDR-p<0.013). The proportion of total effects explained by these significant mediators was between 1.8-22% at age 11, and between 2.9-14% at age 12.

#### Somatoform factor

In unadjusted mediation models of somatoform problems, higher sleep problems, and negative life events, less healthy diet, and lower school involvement were significant mediators of outcomes at age 11 (β between 0.003 and 0.024, FDR-p<0.001). The same variables, in addition to lower parental monitoring, were also significant at age 12 (β between -0.004 and 0.100, FDR-p<0.036). The proportion of total effects explained by these significant mediators was between -0.3 and 21% at age 11 and between -1 and 22% at age 12.

When we adjusted for each mediator and somatoform problems at age 10, mediators showing significant effects on age 11 outcomes in unadjusted models remained significant (β between 0.003 and 0.024, FDR-p<.010). The same mediators, in addition to lower physical activity, were also significant at age 12 (β between 0.003 and 0.016, FDR-p<.021). The proportion of total effects explained by these significant mediators was between 2.4-38% at age 11 and between 1.5-12% at age 12.

#### Sensitivity and additional analyses

When repeating adjusted analyses of age 12 outcomes further controlling for SES and IQ separately, all significant effects remained significant (Tables S4-5), though slightly attenuated. Analyses on the p factor are reported for completeness in Supplement 2 and Tables S6-9, but findings should be interpreted with caution since the p factor included items also included in the neurodevelopmental factor, yielding multicollinearity issues.

## DISCUSSION

This study provides the first robust evidence that a transdiagnostic dimension of neurodevelopmental traits, the neurodevelopmental spectrum^10^, in childhood predicts a broad range of mental health outcomes in early adolescence, and that environmental and lifestyle factors partly underlie these longitudinal associations. In line with our hypotheses, the neurodevelopmental spectrum prospectively predicted major psychopathology dimensions consistent with the HiTOP model, capturing externalizing, internalizing, detachment, and somatoform symptoms. These effects of the neurodevelopmental spectrum emerged over and above baseline mental health difficulties. Among environmental and lifestyle candidate mediators, sleep problems emerged as a key factor underlying these links, as it consistently mediated the associations with all psychopathology dimensions at follow-up, whereas other candidate mediators showed smaller and less consistent mediating effects. Together, our longitudinal findings reveal that sleep problems play a particularly important role in the link between neurodevelopmental traits and a wide range of psychiatric outcomes, highlighting a promising intervention target to improve mental health outcomes in this population.

The identified medium-to-large prospective effects of the neurodevelopmental spectrum at age 10 on later psychiatric dimensions explained a substantial amount of variance (approximately a third) in all outcomes at ages 11 and 12. These longitudinal associations emerged consistently for internalizing and externalizing symptoms, extending previous longitudinal research on depression, anxiety, and behavioral problems in studies of individual NDCs^9,12^. We also report novel prospective associations with detachment and somatoform dimensions, indicating that the neurodevelopmental spectrum predicts early negative psychotic traits that may presage future severe mental illness^21^, as well as poor physical health. Importantly, results were robust to accounting for baseline levels of psychopathology dimensions, meaning that the neurodevelopmental spectrum prospectively predicts within-person change in psychopathology dimensions across early adolescence. By overcoming the limitations of studies based on individual diagnostic categories through a transdiagnostic dimensional approach, these findings indicate that neurodevelopmental traits confer transdiagnostic vulnerability to a broad range of mental health difficulties, rather than showing condition-specific pathways.

Another major contribution of this study is the identification of environmental and lifestyle factors mediating the longitudinal relationship between childhood neurodevelopmental traits and early adolescent psychiatric outcomes. Sleep problems emerged as the strongest and most consistent mediator across all psychiatric dimensions, in both unadjusted models and more stringent adjusted models, with the latter providing stronger evidence of potentially causal pathways^38^. Specifically, we found that higher neurodevelopmental traits predicted worse sleep problems, which in turn predicted higher levels of externalizing, internalizing, detachment, and somatoform symptoms. Effects in adjusted models were small, but emerged consistently across all outcomes. These novel findings extend previous literature on sleep problems in children with ADHD and autism, as well as studies showing that worse sleep is a major risk factor for a variety of negative mental health outcomes^31,32^. Our work suggests that sleep problems are a transdiagnostic lifestyle factor that acts as a significant driver of later psychopathology in children with high neurodevelopmental traits. Possible explanations could be found in the negative impact of increased sensory sensitivity and mind wandering on sleep in youth with high neurodevelopmental traits^45,46^, which in turn can precipitate a variety of mental health problems. Since sleep is modifiable through behavioral and pharmacological interventions, our results indicate that targeting sleep problems may be an effective transdiagnostic intervention approach to mitigate risks for poor mental health outcomes in children with neurodevelopmental traits^47^.

In addition to sleep problems, we found novel evidence that low school involvement partly mediates the developmental association between neurodevelopmental traits and all the examined mental health outcomes, suggesting that fostering inclusion and a sense of belonging in schools might promote positive mental health outcomes in youth with neurodevelopmental traits. Additionally, low levels of parental monitoring and warmth partly underlied the risk of externalizing, internalizing, and detachment symptoms in more stringent adjusted models, with high family conflict similarly impacting internalizing and detachment outcomes. This evidence of mediation builds on previous research showing that parent-child problems are associated with emotional and behavioral problems in ADHD and autism studies^15,23,48^, potentially due to the effects of parenting on emotion dysregulation^49^ and, in turn, mental health outcomes. Similarly, the mediating effects of negative life events on internalizing, detachment, and somatoform outcomes and of neighborhood safety on externalizing and detachment outcomes in adjusted models align with prior research linking traumatic events with poor mental health in youth with NDCs^11,24,25^. With regard to diet, we provide the first evidence for a potential mediating role of unhealthy diet in the link between neurodevelopmental traits and later internalizing, detachment, and somatoform outcomes, building on studies in population samples showing the negative effects of low-quality diet on mental health^34^. Overall, although these findings might suggest a role of parental practices and low-resource environment, it is important to note that these mediators had very small effects in adjusted models. As such, their contribution to poor mental health in this population might be minimal, and interventions targeting these mediators may be unlikely to produce meaningful clinical improvements.

The remaining candidate mediators, namely physical activity and screen time, showed more inconsistent or non-significant effects. While higher levels of physical activity have been associated with better mental health outcomes in population samples^33^, our findings in more stringent adjusted analyses indicate very small significant effects only for somatoform problems at one time point, suggesting a minimal role of physical activity on mental health in youth with neurodevelopmental traits. Regarding screen time, all four types of screen use showed nonsignificant mediating effects when controlling for baseline screen use and mental health, despite showing some significant associations in unadjusted analyses. Despite previous evidence of greater screen use in children with NDCs^50^, our findings show that this is unlikely to lead to poor mental health outcomes in youth with neurodevelopmental traits, in line with findings from strong population-based longitudinal studies^36,37^.

Our findings have significant implications for psychiatric classification and clinical care. This work supports the value of integrating a neurodevelopmental spectrum into transdiagnostic dimensional frameworks, such as HiTOP^21^. Doing so can provide a common classification framework for capturing the co-occurrence and developmental continuity between neurodevelopmental and other psychiatric conditions^10^. This would enhance our ability to identify children at risk for co-occurring presentations and tailor interventions accordingly, before their mental health deteriorates. The strong associations between neurodevelopmental traits and all investigated psychiatric outcomes highlight the importance of regular monitoring of mental health problems in children with neurodevelopmental traits, with an eye to promoting early support. Moreover, the robust transdiagnostic mediation effects of sleep problems suggest that targeted interventions focusing on sleep hygiene could be effective in promoting a wide range of positive mental health outcomes in children with neurodevelopmental traits. Clinicians should routinely assess sleep problems in children with neurodevelopmental traits and consider sleep interventions as part of a comprehensive treatment approach.

Despite the strengths of this study, including our use of a large, socio-demographically diverse longitudinal sample, rigorous statistical analyses, and novel transdiagnostic dimensional approach, several limitations should be noted. First, our analyses included three time points spanning a 3-year period. Although the transition from late childhood to early adolescence is a crucial period for the emergence of mental health problems, particularly in children with NDCs^12^, future studies should replicate our analyses across a longer adolescent follow-up period. Second, our analyses relied on clinical and environmental/lifestyle measures based solely on one reporter (parent or child). Findings should be replicated and extended using multiple reporters (particularly at later adolescent assessments in ABCD) and incorporating objective measures, such as actigraphy and ecological momentary assessment for sleep and other lifestyle factors. Third, while we identified significant mediation effects suggesting potentially causal effects, particularly in longitudinal models adjusting for the respective measures of mediator and outcome variables at age 10, these findings should be triangulated with data from other designs informing on possibly causal links (e.g., randomized controlled trials).

To conclude, this study provides compelling evidence that childhood neurodevelopmental traits are significant predictors of broad psychiatric risk in early adolescence, including externalizing, internalizing, detachment, and somatoform symptoms. The identification of sleep problems as a key mediator, together with small effects of school involvement and family environment, highlights potential intervention targets that could mitigate mental health problems in children with neurodevelopmental traits. These findings underscore the need for transdiagnostic and dimensional approaches in both research and clinical practice, moving beyond categorical diagnoses to better capture the complexity of neurodevelopmental and psychiatric co-occurrence. Future work should continue to explore the mechanisms underlying these associations and develop tailored interventions that address the unique needs of children with neurodevelopmental traits.

## Data Availability

Data used in the preparation of this article were obtained from the Adolescent Brain Cognitive Development (ABCD) Study (https://abcdstudy.org), held in the National Institute of Mental Health Data Archive (NDA). The data are available to authorised researchers with access to the ABCD data.

https://osf.io/yhkvm/

## Acknowledgements

The authors wish to thank all the families who participate in the ABCD Study and all of the ABCD Study staff who make this work possible. We also thank student research assistants Freya Woolford and Alex Oakley for their help preparing supplementary materials. Data used in the preparation of this article were obtained from the Adolescent Brain Cognitive Development (ABCD) Study (https://abcdstudy.org), held in the National Institute of Mental Health Data Archive (NDA). The ABCD Study is supported by the National Institutes of Health (NIH) and additional federal partners under awards numbers U01DA041048, U01DA050989, U01DA051016, U01DA041022, U01DA051018, U01DA051037, U01DA050987, U01DA041174, U01DA041106, U01DA041117, U01DA041028, U01DA041134, U01DA050988, U01DA051039, U01DA041156, U01DA041025, U01DA041120, U01DA051038, U01DA041148, U01DA041093, U01DA041089, U24DA041123, U24DA041147. A full list of supporters is available at https://abcdstudy.org/nih-collaborators. A listing of participating sites and a complete listing of the study investigators can be found at https://abcdstudy.org/principal-investigators.html. ABCD consortium investigators designed and implemented the study and/or provided data but did not necessarily participate in analysis or writing of this report. This manuscript reflects the views of the authors and may not reflect the opinions or views of the NIH or ABCD consortium investigators. The ABCD data repository grows and changes over time. The ABCD data used in this report came from DOI: <u>10.15154/z563-zd24</u>. CC was funded by a London Interdisciplinary Social Science Doctoral Training Partnership (LISS DTP) PhD studentship from the UK Economic and Social Research Council (ESRC). GM was part-funded by a Klingenstein Third Generation Foundation Fellowship (20212999). MM was part-funded by a Jacobs Foundation Research Fellowship. CLM’s research is part-funded by the National Institute for Health and Care Research (NIHR) Maudsley Biomedical Research Centre (BRC).

GM, MM, DR, RB, and CLM served as the statistical experts for this study.

## Presentation information

Preliminary findings from this study were presented as a poster at the 2024 European Network of Hyperkinetic Disorders (EUNETHYDIS) meeting, Cagliari, Italy, October 2024.

## CRediT authorship contribution statement

CC: Conceptualization, Methodology, Formal analysis, Data Curation, Investigation, Visualization, Writing – original draft, Writing – review & editing. DR: Methodology, Data Curation, Writing – review & editing. SL: Data Curation, Writing – review & editing. RB: Methodology, Writing – review & editing. CLM: Methodology, Supervision, Writing – review & editing. MM: Conceptualization, Methodology, Supervision, Writing – review & editing. GM: Conceptualization, Methodology, Investigation, Visualization, Supervision, Funding acquisition, Writing – review & editing.

## References

1. Arora NK, Nair MKC, Gulati S, et al. Neurodevelopmental disorders in children aged 2– 9 years: Population-based burden estimates across five regions in India. PLOS Medicine. 2018;15(7):e1002615. doi:10.1371/journal.pmed.1002615

2. Zablotsky B, Black LI, Maenner MJ, et al. Prevalence and Trends of Developmental Disabilities among Children in the United States: 2009–2017. Pediatrics. 2019;144(4):e20190811. doi:10.1542/peds.2019-0811

3. World Health Organization. International Classification of Diseases. 11th revision. World Health Organization; 2022.

4. American Psychiatric Association. Diagnostic and Statistical Manual of Mental Disorders. 5th ed. American Psychiatric Publishing; 2013.

5. Sokolova E, Oerlemans AM, Rommelse NN, et al. A Causal and Mediation Analysis of the Comorbidity Between Attention Deficit Hyperactivity Disorder (ADHD) and Autism Spectrum Disorder (ASD). J Autism Dev Disord. 2017;47(6):1595–1604. doi:10.1007/s10803-017-3083-7

6. Lonergan A, Doyle C, Cassidy C, et al. A meta-analysis of executive functioning in dyslexia with consideration of the impact of comorbid ADHD. Journal of Cognitive Psychology. 2019;31(7):725–749. doi:10.1080/20445911.2019.1669609

7. Matson JL, Rieske RD, Williams LW. The relationship between autism spectrum disorders and attention-deficit/hyperactivity disorder: An overview. Research in Developmental Disabilities. 2013;34(9):2475–2484. doi:10.1016/j.ridd.2013.05.021

8. Gidziela A, Ahmadzadeh YI, Michelini G, et al. A meta-analysis of genetic effects associated with neurodevelopmental disorders and co-occurring conditions. Nat Hum Behav. 2023;7(4):642–656. doi:10.1038/s41562-023-01530-y

9. Augustine L, Lygnegård F, Granlund M. Trajectories of participation, mental health, and mental health problems in adolescents with self-reported neurodevelopmental disorders. Disabil Rehabil. 2022;44(9):1595–1608. doi:10.1080/09638288.2021.1955304

10. Michelini G, Carlisi CO, Eaton NR, et al. Where do neurodevelopmental conditions fit in transdiagnostic psychiatric frameworks? Incorporating a new neurodevelopmental spectrum. World Psychiatry. 2024;23(3):333–357. doi:10.1002/wps.21225

11. Dardani C, Schalbroeck R, Madley-Dowd P, et al. Childhood Trauma As a Mediator of the Association Between Autistic Traits and Psychotic Experiences: Evidence From the Avon Longitudinal Study of Parents and Children Cohort. Schizophr Bull. 2023;49(2):364–374. doi:10.1093/schbul/sbac167

12. Thapar A, Livingston LA, Eyre O, Riglin L. Practitioner Review: Attention-deficit hyperactivity disorder and autism spectrum disorder – the importance of depression. Journal of Child Psychology and Psychiatry. 2023;64(1):4–15. doi:10.1111/jcpp.13678

13. Embracing Complexity Research Priority Setting. Top 10 Priorities for Research on Neurodivergence. 2024. https://www.autistica.org.uk/downloads/files/Embracing-Complexity-2024_2024-03-14-155013_wvts.pdf

14. Benevides TW, Shore SM, Palmer K, et al. Listening to the autistic voice: Mental health priorities to guide research and practice in autism from a stakeholder-driven project. Autism. 2020;24(4):822–833. doi:10.1177/1362361320908410

15. Humphreys KL, Katz SJ, Lee SS, Hammen C, Brennan PA, Najman JM. The association of ADHD and depression: mediation by peer problems and parent-child difficulties in two complementary samples. J Abnorm Psychol. 2013;122(3):854–867. doi:10.1037/a0033895

16. Carter Leno V, Hollocks MJ, Chandler S, et al. Homotypic and Heterotypic Continuity in Psychiatric Symptoms From Childhood to Adolescence in Autistic Youth. J Am Acad Child Adolesc Psychiatry. 2022;61(12):1445–1454. doi:10.1016/j.jaac.2022.05.010

17. Einfeld SL, Ellis LA, Emerson E. Comorbidity of intellectual disability and mental disorder in children and adolescents: a systematic review. J Intellect Dev Disabil. 2011;36(2):137–143. doi:10.1080/13668250.2011.572548

18. Vos M, Wang R, Rommelse NNJ, Snieder H, Larsson H, Hartman CA. Familial co-aggregation and shared familiality among neurodevelopmental problems and with aggressive behavior, depression, anxiety, and substance use. Psychological Medicine. 2024:1–13. doi:10.1017/S003329172400309X

19. Astle DE, Holmes J, Kievit R, Gathercole SE. Annual Research Review: The transdiagnostic revolution in neurodevelopmental disorders. Journal of Child Psychology and Psychiatry. 2022;63(4):397–417. doi:10.1111/jcpp.13481

20. Michelini G, Barch DM, Tian Y, Watson D, Klein DN, Kotov R. Delineating and validating higher-order dimensions of psychopathology in the Adolescent Brain Cognitive Development (ABCD) study. Transl Psychiatry. 2019;9(1):1–15. doi:10.1038/s41398-019-0593-4

21. Kotov R, Krueger RF, Watson D, et al. The Hierarchical Taxonomy of Psychopathology (HiTOP): A dimensional alternative to traditional nosologies. Journal of Abnormal Psychology. 2017;126(4):454–477. doi:10.1037/abn0000258

22. Powell V, Riglin L, Hammerton G, et al. What explains the link between childhood ADHD and adolescent depression? Investigating the role of peer relationships and academic attainment. Eur Child Adolesc Psychiatry. 2020;29(11):1581–1591. doi:10.1007/s00787-019-01463-w

23. Song J, Fogarty K, Suk R, Gillen M. Behavioral and mental health problems in adolescents with ADHD: Exploring the role of family resilience. Journal of Affective Disorders. 2021;294:450–458. doi:10.1016/j.jad.2021.07.073

24. Rychik N, Fassett-Carman A, Snyder HR. Dependent Stress Mediates the Relation Between ADHD Symptoms and Depression. J Atten Disord. 2021;25(12):1676–1686. doi:10.1177/1087054720925900

25. Rai D, Culpin I, Heuvelman H, et al. Association of Autistic Traits With Depression From Childhood to Age 18 Years. JAMA Psychiatry. 2018;75(8):835–843. doi:10.1001/jamapsychiatry.2018.1323

26. Del Bianco T, Lockwood Estrin G, Tillmann J, et al. Mapping the link between socio-economic factors, autistic traits and mental health across different settings. Autism. 2024;28(5):1280–1296. doi:10.1177/13623613231200297

27. Mori H, Hirota T, Monden R, Takahashi M, Adachi M, Nakamura K. School Social Capital Mediates Associations Between ASD Traits and Depression Among Adolescents in General Population. J Autism Dev Disord. 2023;53(10):3825–3834. doi:10.1007/s10803-022-05687-9

28. Butler AM, Kowalkowski M, Jones HA, Raphael JL. The Relationship of Reported Neighborhood Conditions With Child Mental Health. Academic Pediatrics. 2012;12(6):523–531. doi:10.1016/j.acap.2012.06.005

29. Yu X, Rahman MM, Carter SA, et al. Neighborhood Disadvantage and Autism Spectrum Disorder in a Population With Health Insurance. JAMA Psychiatry. 2024;81(2):209–213. doi:10.1001/jamapsychiatry.2023.4347

30. Breedvelt JJF, Tiemeier H, Sharples E, et al. The effects of neighbourhood social cohesion on preventing depression and anxiety among adolescents and young adults: rapid review. BJPsych Open. 2022;8(4):e97. doi:10.1192/bjo.2022.57

31. Dimakos J, Gauthier-Gagné G, Lin L, Scholes S, Gruber R. The Associations Between Sleep and Externalizing and Internalizing Problems in Children and Adolescents with Attention-Deficit/Hyperactivity Disorder: Empirical Findings, Clinical Implications, and Future Research Directions. Psychiatr Clin North Am. 2024;47(1):179–197. doi:10.1016/j.psc.2023.06.012

32. Fletcher FE, Foster-Owens MD, Conduit R, Rinehart NJ, Riby DM, Cornish KM. The developmental trajectory of parent-report and objective sleep profiles in autism spectrum disorder: Associations with anxiety and bedtime routines. Autism. 2017;21(4):493–503. doi:10.1177/1362361316653365

33. Rodriguez-Ayllon M, Cadenas-Sánchez C, Estévez-López F, et al. Role of Physical Activity and Sedentary Behavior in the Mental Health of Preschoolers, Children and Adolescents: A Systematic Review and Meta-Analysis. Sports Med. 2019;49(9):1383–1410. doi:10.1007/s40279-019-01099-5

34. Khalid S, Williams CM, Reynolds SA. Is there an association between diet and depression in children and adolescents? A systematic review. British Journal of Nutrition. 2016;116(12):2097–2108. doi:10.1017/S0007114516004359

35. Kozyra M, Zimnicki P, Kaczerska J, Śmiech N, Nowińska M, Milanowska J. Relationship between the diet and psychiatric diseases such as depression, anxiety and attention-deficit hyperactivity disorder (ADHD). *Journal of Education*, Health and Sport. 2020;10(8):93–104. doi:10.12775/JEHS.2020.10.08.011

36. Nagata JM, Al-Shoaibi AAA, Leong AW, et al. Screen time and mental health: a prospective analysis of the Adolescent Brain Cognitive Development (ABCD) Study. BMC Public Health. 2024;24(1):2686. doi:10.1186/s12889-024-20102-x

37. Tang S, Werner-Seidler A, Torok M, Mackinnon AJ, Christensen H. The relationship between screen time and mental health in young people: A systematic review of longitudinal studies. Clin Psychol Rev. 2021;86:102021. doi:10.1016/j.cpr.2021.102021

38. Loh WW, Ren D. Adjusting for Baseline Measurements of the Mediators and Outcome as a First Step Toward Eliminating Confounding Biases in Mediation Analysis. Perspect Psychol Sci. 2023;18(5):1254–1266. doi:10.1177/17456916221134573

39. Garavan H, Bartsch H, Conway K, et al. Recruiting the ABCD sample: Design considerations and procedures. Dev Cogn Neurosci. 2018;32:16–22. doi:10.1016/j.dcn.2018.04.004

40. Achenbach TM. Manual for ASEBA School-Age Forms & Profiles. University of Vermont, Research Center for Children, Youth & Families. 2001.

41. Muthén B, Muthén L. Mplus. In: Handbook of Item Response Theory. Chapman and Hall/CRC; 2017.

42. Müller S, Scealy JL, Welsh AH. Model Selection in Linear Mixed Models. Statistical Science. 2013;28(2):135–167. doi:10.1214/12-STS410

43. Benjamini Y, Hochberg Y. Controlling the False Discovery Rate: A Practical and Powerful Approach to Multiple Testing. Journal of the Royal Statistical Society: Series B (Methodological). 1995;57(1):289–300. doi:10.1111/j.2517-6161.1995.tb02031.x

44. Wechsler D. Wechsler Intelligence Scale for Children; Manual. The Psychological Corp. 1949.

45. Fell J. Mind wandering, poor sleep, and negative affect: a threefold vicious cycle? Front Hum Neurosci. 2024;18:1441565. doi:10.3389/fnhum.2024.1441565

46. Tzischinsky O, Meiri G, Manelis L, et al. Sleep disturbances are associated with specific sensory sensitivities in children with autism. Molecular Autism. 2018;9(1):22. doi:10.1186/s13229-018-0206-8

47. Papadopoulos N, Sciberras E, Hiscock H, Mulraney M, McGillivray J, Rinehart N. The Efficacy of a Brief Behavioral Sleep Intervention in School-Aged Children With ADHD and Comorbid Autism Spectrum Disorder. J Atten Disord. 2019;23(4):341–350. doi:10.1177/1087054714568565

48. Tarver J, Palmer M, Webb S, et al. Child and parent outcomes following parent interventions for child emotional and behavioral problems in autism spectrum disorders: A systematic review and meta-analysis. Autism. 2019;23(7):1630–1644. doi:10.1177/1362361319830042

49. Evans S, Bhide S, Quek J, et al. Mindful Parenting Behaviors and Emotional Self-Regulation in Children With ADHD and Controls. Journal of Pediatric Psychology. 2020;45(9):1074–1083. doi:10.1093/jpepsy/jsaa073

50. Yuan G, Zhu Z, Guo H, et al. Screen Time and Autism Spectrum Disorder: A Comprehensive Systematic Review of Risk, Usage, and Addiction. J Autism Dev Disord. Published online December 3, 2024. doi:10.1007/s10803-024-06665-z

