## Supplementary material for "Developmental pathways from childhood neurodevelopmental traits to early adolescent psychiatric dimensions: the role of environmental and lifestyle factors": Caserini_SupplMaterials.docx

**SUPPLEMENTAL MATERIALS**

**Supplement 1 - Supplemental methods**

Our analyses included a wide range of environmental and lifestyle factors hypothesized to mediate the link between neurodevelopmental traits and later mental health outcomes.

Environmental factors included:

1. *Parental warmth* was measured with the Children’s Report of Parent Behavior Inventory (CRPBI)^1^. The ABCD study only included a short version of the original Acceptance Scale (5 items), which evaluates the child’s perceptions of parent warmth, acceptance, and responsiveness on a 3-point scale.
2. *Parental monitoring* was measured using the Parental Monitoring Survey, a self-administrated child-report survey of 5 items investigating the children’s perception of their parent’s vigilance when they are at home and when they are outside on a 5-point scale^2–4^. In the current study, we used the Parental Monitoring Survey summary score.
3. *Family conflict* was measured using the parent-report Family Conflict subscale from the Family Environment Scale (FES)^5^, a 90-item self-report questionnaire that investigates the social and environmental climate in the family, primarily addressing the interpersonal relationship between family members, personal growth, and system maintenance. The Family Conflict Subscale (9 items) explores conflicts among family members on a 2-point scale.
4. *School involvement* was measured with the School Involvement Subscale from the School Risk and Protective Factors Protocol (SRPF), a child-report survey derived from the Communities That Care (CTC) Youth Survey^6^. The School Involvement subscale (4 items) explores the extent to which the child feels involved and comfortable in the school activities on a 4-point scale.
5. *Negative life events* were measured with the child-report Adverse Life Events Scale^7^, a 25-item questionnaire that investigates a variety of experiences over which children have little or no control. Children were asked to report whether they had ever experienced a given event and rate whether it had been “mostly good” or “mostly bad” for them. Example items include: “Someone in family died” and “Was a victim of crime/violence/assault”. In the current project, we used a sum score of lifetime events rated as negative; if a child rated no lifetime event as negative, their score was 0.
6. *Neighborhood safety* was measured with the Neighborhood Safety/Crime Survey, child-report 1-item questionnaire from the PhenX Toolkit’s Neighborhood Safety Protocol^8^. This measure investigates children’s feelings on whether their neighborhood is safe from crime, on a 5-point scale. Main analyses on neighborhood safety used this self-report measure, for consistency with the other self-reported environmental mediators and because a child’s neurodevelopmental traits may be more likely to influence their own perception of safety, as opposed to their parents’ perceptions. As the self-report version included only one item, additional analyses were run on the equivalent parent-reported measure of perceived neighborhood safety, including three items about safety and the presence of crime in the neighborhood (Table S10).

Lifestyle factors included:

1. *Sleep problems* were assessed from the parent-report summary score of the Sleep Disturbance Scale for Children (SDSC)^9^, a 27-item questionnaire designed to measure sleep problems in children and adolescents. The response is provided on a 5- point scale investigating the frequency of sleep disturbance experienced by the child in the past 6 months, with higher scores indicating worse sleep.
2. *Screen usage* was measured through the Youth Self-report Screen Time Questionnaire^10^, a survey that explores the average daily time spent on screen devices during weekdays and weekends. Answers have been coded on a 7 points scale: 0 = None; .25 = < 30 minutes; 0.5 = 30 minutes; 1 = 1 hour; 2 = 2 hours; 3 = 3 hours; 4 = 4+ hours. For this study, average daily use across the week was calculated based on average weekday and weekend use. We created four sub-composites to explore how specific types of screen use may affect mental health, informed by correlations between items. *Passive watching* captured the amount of time watching television or movies. *Active gaming* captured the amount of time playing video games on a computer or other devices. Social engagement captured the amount of time spent texting or video-calling on a cellphone, tablet, or computer. *Social media* captured the amount of time spent on social networking sites. Correlations between the four screen sub-composites were small to medium (from .17 to .46), confirming that they captured relatively distinct aspects of screen use.
3. *Diet* was measured through parent-report questions investigating the nutrition patterns of the child in the past year. In the present study, we used the summary score of 14 yes/no items about healthy diet, with higher scores reflecting healthier dietary habits.
4. *Physical activity* was measured using The Sports and Activities Involvement Questionnaire (SAI-Q), parent-report version^11^. The SAI-Q measured lifetime and past year involvement in 23 different sports, as well as frequency and duration. In this study, we considered the items investigating the frequency of specific physical activity per week. For each specific sport item investigating frequency (e.g., “Field Hockey: About how many days per week?”) the original responses were: 0 = 0 day per week; 1 = 1 day per week; 2 = 2 days per week; 3 = 3 days per week; 4 = 4 days per week; 5 = 5 days per week; 6 = 6 days per week; 7 = 7 days per week; 8 = Once every 2 weeks; 9 = One day every month; 10 = Less than one day per month; 999 = Don't know. We recoded these values as follows: 0 = 0 (0 day per week); 1 = 4 (1 day per week); 2 = 5 (2 days per week); 3 = 6 (3 days per week); 4 = 7 (4 days per week); 5 = 8 (5 days per week); 6 = 9 (6 days per week); 7 = 10 (7 days per week); 8 = 3 (once every 2 weeks); 9 = 2 (one day every month); 10 = 1 (less than one day every month); 999 = NA (missing value). We then aggregated the recoded items into a sum score for sports activity, estimating the total amount of sport practiced in a typical week.

**Supplement 2 – Supplemental Results**

*Longitudinal associations between neurodevelopmental factor and p factor*

In unadjusted models, the neurodevelopmental factor at baseline significantly predicted the p factor at age 11 and 12 (β between .694 and .635, p<.001), explaining up to 48% of the variance (Table S6). Effects were reduced but remained significant in adjusted models at age 11 (β=.046, p=.004) controlling for p at age 10, whereas the association at age 12 became non-significant (β=.020, p=.264) (Table S6).

*Mediation models with p factor as outcome*

In primary mediation models of the p factor, all environmental and lifestyle factors were significant mediators of age-11 outcomes (β between .002 and .125, FDR-p<.008), except for physical activity, social media, and social engagement (Table S7). Sleep, diet, family conflict, parental monitoring, parental warmth, negative life events, neighborhood safety, and school involvement were significant mediators in analyses of age-12 outcomes (β between .002 and .094, FDR-p<.001) (Table S7). The proportion of total effects explained by these significant mediators was between 0.3% and 17% for age-11 p factor, and between 0.4% and 14% for age-12 p factor.

In adjusted models controlling for each mediator and p factor at age 10, diet, parental monitoring, parental warmth, negative life events, and school involvement were significant in age-11 models (β between -.006 and .009, FDR-p<.016) (Table S8). Diet, parental monitoring, parental warmth, and school involvement were significant mediators in analyses of age-12 models (β between -.005 and .007, FDR-p<.033) (Table S8). However, these models did not converge, as indicated by issues in computing reasonable proportions of total effect mediated and 95% confidence interval estimates through bootstrapping, thus these results should be interpreted cautiously.

Results were unchanged when accounting for socioeconomic status and IQ (Table S9).

**Supplemental Reference List**

1.	Schaefer ES. Children’s Reports of Parental Behavior: An Inventory. *Child Development*. 1965;36(2):413-424. doi:10.2307/1126465

2.	Karoly HC, Callahan T, Schmiege SJ, Ewing SWF. Evaluating the Hispanic Paradox in the Context of Adolescent Risky Sexual Behavior: The Role of Parent Monitoring. *J Pediatr Psychol*. 2016;41(4):429-440. doi:10.1093/jpepsy/jsv039

3.	Shillington AM, Lehman S, Clapp J, Hovell MF, Sipan C, Blumberg EJ. Parental monitoring: Can it continue to be protective among high-risk adolescents? *Journal of Child & Adolescent Substance Abuse*. 2005;15(1):1-15. doi:10.1300/J029v15n01_01

4.	DiClemente RJ, Wingood GM, Crosby R, et al. Parental monitoring: association with adolescents’ risk behaviors. *Pediatrics*. 2001;107(6):1363-1368. doi:10.1542/peds.107.6.1363

5.	Moos, R. H. *Family Environment Scale Manual: Development, Applications, Research*. Consult. Psychol. Press; 1994.

6.	Arthur MW, Briney JS, Hawkins JD, Abbott RD, Brooke-Weiss BL, Catalano RF. Measuring risk and protection in communities using the Communities That Care Youth Survey. *Evaluation and Program Planning*. 2007;30(2):197-211. doi:10.1016/j.evalprogplan.2007.01.009

7.	Tiet QQ, Bird HR, Davies M, et al. Adverse Life Events and Resilience. *Journal of the American Academy of Child & Adolescent Psychiatry*. 1998;37(11):1191-1200. doi:10.1097/00004583-199811000-00020

8.	Mujahid MS, Diez Roux AV, Morenoff JD, Raghunathan T. Assessing the Measurement Properties of Neighborhood Scales: From Psychometrics to Ecometrics. *American Journal of Epidemiology*. 2007;165(8):858-867. doi:10.1093/aje/kwm040

9.	Bruni O, Ottaviano S, Guidetti V, et al. The Sleep Disturbance Scale for Children (SDSC) Construction and validation of an instrument to evaluate sleep disturbances in childhood and adolescence. *Journal of Sleep Research*. 1996;5(4):251-261. doi:10.1111/j.1365-2869.1996.00251.x

10.	Bagot K, Tomko R, Marshall AT, et al. Youth screen use in the ABCD® study. *Developmental Cognitive Neuroscience*. 2022;57:101150. doi:10.1016/j.dcn.2022.101150

11.	Barch DM, Albaugh MD, Avenevoli S, et al. Demographic, physical and mental health assessments in the adolescent brain and cognitive development study: Rationale and description. *Dev Cogn Neurosci*. 2018;32:55-66. doi:10.1016/j.dcn.2017.10.010
